# The qMini assay identifies an overlooked class of splice variants

**DOI:** 10.1101/2023.11.02.23297963

**Authors:** Bin Guan, Chelsea Bender, Madhulatha Pantrangi, Nia Moore, Melissa Reeves, Amelia Naik, Huirong Li, Kerry Goetz, Delphine Blain, Aime Agather, Catherine Cukras, Wadih M. Zein, Laryssa A. Huryn, Brian P. Brooks, Robert B. Hufnagel

**Affiliations:** Ophthalmic Genetics and Visual Function Branch, National Eye Institute, National Institutes of Health, Bethesda, MD; Department of Pathology and Laboratory Medicine, Weill Cornell Medicine, New York, NY; Office of Data Science and Health Informatics, National Eye Institute, National Institutes of Health, Bethesda, MD

**Author notes:** **Corresponding authors:** &.

## Abstract

Splice variants are known to cause diseases by utilizing alternative splice sites, potentially resulting in protein truncation or mRNA degradation by nonsense-mediated decay. Splice variants are verified when altered mature mRNA sequences are identified in RNA analyses or minigene assays. Using a quantitative minigene assay, qMini, we uncovered a previously overlooked class of disease-associated splice variants that did not alter mRNA sequence but decreased mature mRNA level, suggesting a potentially new pathogenic mechanism.

## Main text

In the era of personalized genomic medicine, disease-associated splicing variants are typically identified from DNA sequencing aided by *in silico* prediction tools. If patient RNA is available, RNA analysis is used to verify whether a candidate variant causes a splicing defect, i.e., mRNA sequence alteration. However, patient RNA samples, particularly those from relevant tissues, are often unavailable in clinical testing. In addition, a patient RNA sample alone is insufficient to confirm the causative relationship between a candidate splicing variant and an aberrant splicing event, as the result reflects a broader genetic and physiologic background such as other variants *in cis* or *in trans* and epigenetic effects^1^. Thus, *in vitro* splicing assays, such as minigene or midigene, are widely used to demonstrate whether a variant causes aberrant splicing by defining the resulting sequence alteration in mature mRNA^2^. As such, the assay typically involves the construction of two plasmids differing only at the variant site, transfection of the plasmids into a cell line, reverse-transcription PCR (RT-PCR), gel-electrophoresis, and sequencing.

We selected 15 candidate noncanonical splice variants within 60 nucleotides from the canonical splice sites from individuals suspected to have an inherited eye disease for minigene assays (Supplementary Table 1). Since two of these variants *RPE65*:c.11+5G>A and *RS1*:c.52+5G>C are located in the first intron of their respected genes, genomic DNA sequence from *RPE65* exon 1 to part of intron 2 and DNA sequence from *RS1* exon 1 and part of intron 1 were cloned into the RHCglo vector^3^, so that exon 1 remains as the first exon in the minigene constructs (Fig. 1a, Online Methods). The other 13 variants are located in the middle of the gene, thus their corresponding genomic DNA fragments were cloned to replace the middle exon in the RHCglo vector (Fig. 1a). Eight variants caused splicing alteration detectable by gel-electrophoresis and confirmed by Sanger sequencing. The remaining seven variants showed the same migration patterns on the gel as their reference constructs, and Sanger sequencing confirmed that there was no sequence alteration present in the PCR products (Fig. 1b-f and Supplementary Tables 1 & 2). In addition to the 15 noncanonical splice variants, two canonical splice site variants in the *RS1* gene, c.52+1G>A and c.52+1G>T, were also included and faint bands corresponding to the mature mRNA were barely detectable on the gel image (Fig. 1c), suggesting that splicing occurred but at a very low efficiency.

**Fig. 1.**
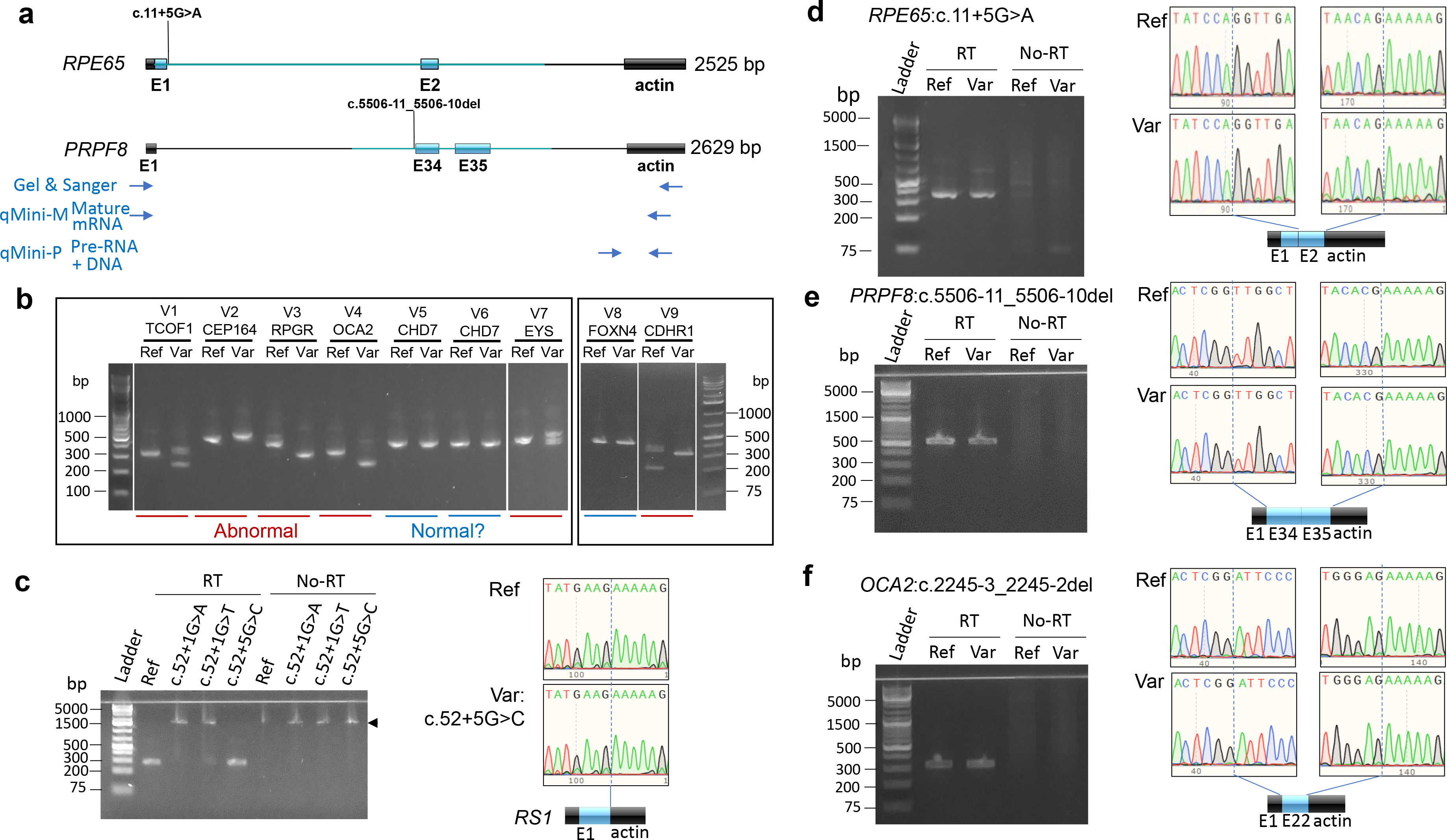
Disease-associated variants may not lead to mature mRNA sequence changes in minigene assay. **a)** Diagram of minigene constructs in the RHCglo vector. The primer binding positions are shown for Sanger sequencing PCR and qPCR or dPCR to detect mature mRNA (qMini-M) or pre-mRNA/DNA (qMini-P). Boxes indicate exon and lines indicate intron. Black and blue boxes or lines represent vector and human DNA sequences, respectively. **b)** Agarose gel electrophoresis images of reverse transcription-PCR (RT-PCR) products of nine candidate splicing variants from the minigene assay. Detailed variant information is listed in Supplementary Table 1. **c-f)** Agarose gel electrophoresis images and Sanger chromatograms of RT-PCR products from minigene constructs showing four noncanonical splice variants not causing mRNA sequence alteration. The blue dashed lines on the chromatograms mark the exon-exon junctions in the minigene cDNA. Arrowhead, product from plasmid DNA or precursor mRNA.

We hypothesized that some splicing variants could alter mature mRNA levels by affecting splicing efficiency, which could be masked by end-point PCR reactions. Thus, we attempted quantitative PCR (qPCR) to measure the steady-state levels of mature mRNA using the primer pair qMini-M (Fig. 1a) in samples from the seven noncanonical variants that did show mRNA sequence alteration. Interestingly, the mature mRNA levels of three variants, *RPE65*:c.11+5G>A, *RS1*:c.52+5G>C, and *PRPF8*:c.5506-11_5506-10del were reduced 667, 4.1, and 5.1 folds respectively in the variant samples as compared to reference samples, while those of the other four did not change (Fig. 2a & Supplementary Table 1).

**Fig. 2.**
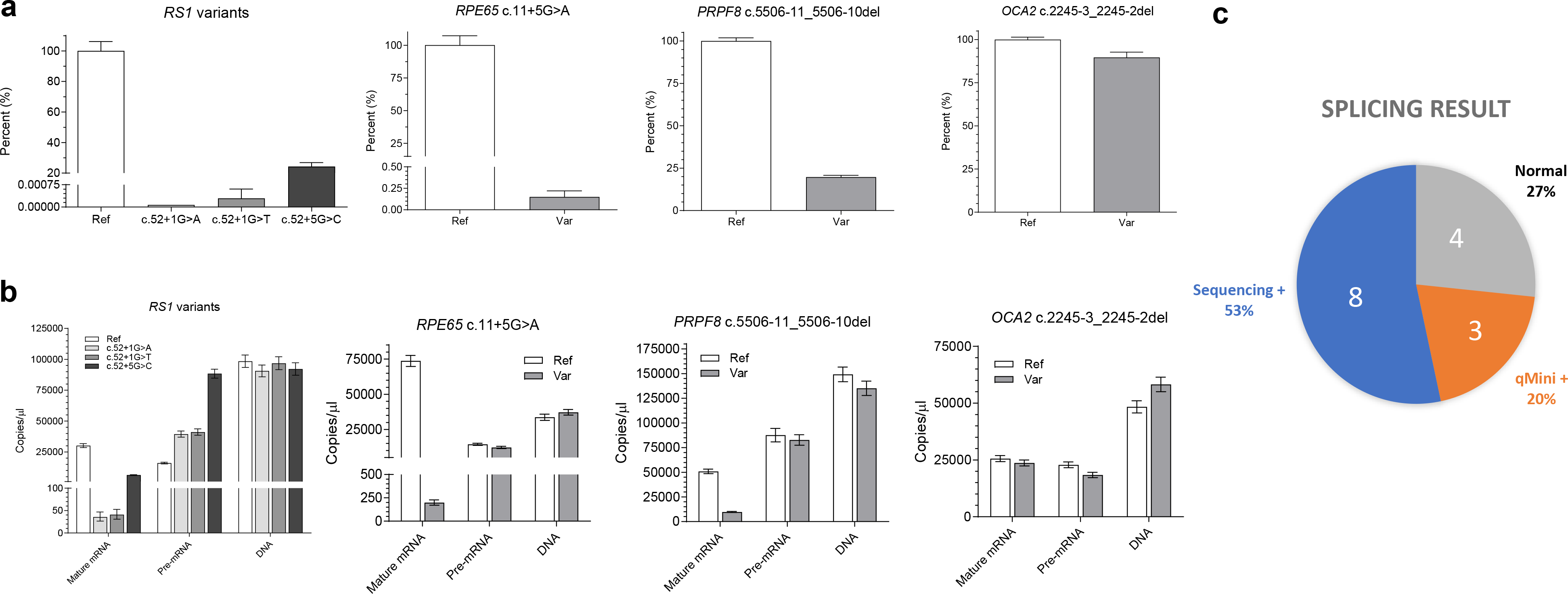
Candidate splice variants causing diminished mature mRNA levels. **a)** Quantitative PCR (qPCR) of minigene assay cDNA quantifying the mature minigene mRNA after normalization to the *RPP30* gene. Error bar, standard deviation. **b)** Digital droplet PCR (dPCR) of minigene assay cDNA quantifying the mature minigene mRNA, pre-mRNA, and plasmid DNA present in the RNA samples. Error bar, Poisson 95% confidence intervals. **c)** Pie chart of the splicing result for the 15 noncanonical variants.

The minigene expression is driven by the RSV promoter, whose activity is not expected to be influenced by variants introduced in the constructs. The steady state mature-to-precursor (M/P) ratio of mRNA was found to correlate with the rate constant of the splicing reaction^4^. Thus, we employed digital droplet PCR (dPCR) to quantify the steady-state levels of plasmid DNA, pre-mRNA, and mature RNA in the RNA samples to assess splicing efficiencies. The primer pair qMini-P amplifies both the DNA and cDNA derived from the pre-mRNA (Fig. 1a). The pre-mRNA levels were obtained by subtracting the plasmid DNA amounts measured in no-reverse transcriptase controls from cDNA reactions. The pre-mRNA levels were similar between the ref-var pairs for *RPE65*:c.11+5G>A, *PRPF8*:c.5506-11_5506-10del and *OCA2*:c.2245-3_2245-2del. In contrast, the pre-mRNA levels in the *RS1*:c.52+1G>A and c.52+1G>T samples increased ∼2.5 folds as compared to the reference construct, and pre-mRNA in the c.52+5G>C sample increased 5.5 folds. Intriguingly, the pre-mRNA/DNA ratios of the reference constructs varied from 0.16-0.59 and the pre-mRNA/DNA ratios, when comparing variant to reference constructs, either increased, decreased, or remained the same: *RS1* ref:c.52+1G>A:c.52+1G>T:c.52+5G>C = 0.16:0.44:0.42:0.96, *RPE65* ref:var = 0.43:0.33, *OCA2* ref:var = 0.47:0.32, and *PRPF8* ref:var = 0.59:0.61. These data suggest that the pre-mRNA steady-state levels may be influenced by DNA sequence, transcription, splicing efficiency, or other factors. Nevertheless, showing equivalent input amounts of the ref:var plasmid DNA serves as an internal control for the qMini assay. After normalizing DNA inputs, dPCR showed that the mature mRNA of *RS1*:c.52+5G>C variant decreased 4.5 folds as compared to the reference construct, *PRPF8*:c.5506-11_5506-10del decreased 4.7 folds, and *RPE65*:c.11+5G>A decreased 408 folds, which were consistent with the qPCR data. Examination of the M/P ratios showed that the variants *RS1*:c.52+5G>C, *PRPF8*:c.5506-11_5506-10del, and *RPE65*:c.11+5G>A decreased splicing efficiency by 27, 4.9, and 312 folds, respectively, as compared to their corresponding constructs with reference sequences. The M/P ratios for the *OCA2* reference sequence and c.2245-3_2245-2del variant are similar at 1.1 and 1.3, respectively.

The ClinGen SVI Splicing Subgroup recently provided guidelines on splicing variant interpretation and recommended applying the strong benign criterion BP7_S if no variant-specific impact observed in RNA/splicing data^5^. Our data indicate that BP7_S should not be applied until the qMini result is found to be negative, as assigning BP7_S to a rare variant (PM2, rare in population databases) would have led it to be classified as likely benign according the Bayesian classification approach^6^. Consideration of qMini allows us to reclassify the three qMini-positive variants, *PRPF8*:c.5506-11_5506-10del from variant of uncertain significance (VUS) to likely pathogenic, *RS1*:c.52+5G>C from VUS to pathogenic, and *RPE65*:c.11+5G>A from likely pathogenic to pathogenic (Online Discussion)^5^. Applying the BP7_S to the four qMini-negative splice variants led us to reclassify them to likely benign from VUS; of note, individual NEI-3 was subsequently found to harbor another pathogenic variant in another gene that is consistent with his presentation (Supplementary Table 1).

In summary, the qMini assay is a robust assay to identify candidate variants that alter mature mRNA levels and splicing efficiency while using canonical splice sites. These variants represent a largely overlooked class of splicing variants associated with genetic diseases (Extended Date Fig. 1). In this limited dataset of 15 noncanonical splice variants, 20% (3/15) were conventional-minigene-negative but qMini-positive (Fig. 2c). Thus, this qMini assay could improve the clinical validity and utility of NGS tests by establishing the pathogenicity of variants that initially classified as benign based on the absence of mRNA sequence changes. Two of the three qMini-positive variants tested here had low SpliceAI and Pangolin scores (Supplementary Table 1), suggesting that building a larger category of qMini-positive variants could facilitate the development of new splice prediction tools and shed new light on their pathomechanisms.

## Online Discussion

The *RS1*:c.52+5G>C variant has been found *in cis* to the c.35T>A p.(Leu12His) variant in multiple individuals with X-linked retinoschisis^7, 8^. Pathogenicity of the c.52+5G>C variant was unclear because *in silico* prediction tools predict it to be tolerated^8^. The L12H variant prevents signal peptide cleavage, leading to impaired translocation to endoplasmic reticulum and proteasomal degradation^9^. Our data indicate that the c.52+5G>C variant is also pathogenic due to diminished splicing efficiency per the qMini assay.

The *RPE65*:c.11+5G>A variant has been observed in at least five probands in the homozygous state and in at least 27 probands in the heterozygous state together with another *RPE65* pathogenic variant in the literature (Supplementary Table 3). A recent study applied the midigene approach and found that the variant produces a minor mRNA species with a 124-nt intron retention and a majority of mRNA from normal splicing^10^. Interestingly, *RPE65* mRNA and protein levels are markedly diminished in patient-derived retinal pigment epithelial (RPE) cells, which was not rescued by inhibition of nonsense-mediated decay, leading to the conclusion that the variant causes diminished *RPE65* expression independent of splicing^10^. Our *in vitro* qMini assay data supports the conclusion that the variant leads to a lower splicing efficiency and subsequently diminished mRNA and protein levels in the patient derived RPE cells.

The *PRPF8*:c.5506-11_5506-10del variant was found in a sporadic individual diagnosed with rod cone dystrophy (Extended Data Fig. 2). It deletes a TT (or UU) dinucleotide in the polypyrimidine tract that has been relatively evolutionarily conserved among vertebrates (Extended Data Fig. 3). Showing diminished mature RNA in the qMini assay allowed us to apply the strong pathogenic supporting criterion PVS1_S for variant classification, leading us to classify the variant as likely pathogenic according to the ACMG/AMP sequence variant classification guideline and a recent recommendation on splicing variant classification from the ClinGen SVI Splicing Subgroup (PVS1_S; PM2, absent in gnomAD)^5, 11^. The polypyrimidine tract is a critical splicing element located between an acceptor site and the lariat branchpoint, which aids the recognition and binding of U2 auxiliary factor to the 3’ splice site^12^. Shortening of the polypyrimidine tract by the c.5506-11_5506-10del variant may be the underlying mechanism of diminished mature mRNA. It is notable that another polypyrimidine variant *OCA2*:NM_000275.3:c.1045-9T>G tested in this study led to the skipping of the nearby exon 10 as well as a 4-fold reduction of mature mRNA (Supplementary Table 1 and data not shown).

The *OCA2* exon 10 contains the complete triplet codons coding for a.a. Ile349 to Asp372, thus preserving the reading frames in the minigene mature mRNA. Additional studies would be required to understand why polypyrimidine tract variants lead to different splicing outcomes, although it is conceivable that the local DNA context plays a role.

## Online Methods

### Human subjects and genetic testing

Ten variants from ten individuals evaluated in the NEI eye clinic were included in the study (Subject ID, NEI-1 to −10 in Supplementary Table 1). Human samples and data were collected under research protocols approved by the NIH Institutional Review Board following Federalwide Assurance principles. NGS-panel or exome/genome testing was performed either in house or via a commercial diagnostic laboratory. Three variants (Subject ID PRG-1 to −3 in Supplementary Table 1) were from a consult service for a commercial diagnostic company. Individuals with two *PNPLA6* variants V14 and V15 (Supplementary Table 1) were recently published^13^.

### DNA constructs

The plasmid RHCglo, a gift from Thomas Cooper (Addgene plasmid # 80169)^3^, was used for cloning of minigene constructs. A majority of the minigene plasmids except *RPE65* and *RS1* were made by gene synthesis and mutagenesis (LifeSct, Rockville, MD). *RS1* constructs were made by PCR amplification of the exon 1 and part of intron 1 of *RS1* gene using the patient DNA with or without variants as templates, which were then cloned into the BspEI and SalI sites of the RHCglo vector using the NEBuilder HiFi DNA assembly kit (NEB). To make the *RPE65* constructs using the HiFi assembly kit, part of the *RPE65* exon 1 to intron 2 sequence was amplified from a human genomic DNA sample and was cloned into the BspEI and XbaI sites within the RHCglo plasmid. The variant c.11+5G>A was introduced into the construct during the PCR and HiFi assembly steps. The genomic coordinates of the cloned human DNA fragments are listed in Supplementary Table 1. All constructs were verified by Sanger sequencing.

### Variant splicing effect prediction

The SpliceAI and Pangolin predictions were gathered from the SpliceAI-lookup website (https://spliceailookup.broadinstitute.org accessed on 10/23/2023). The SpliceAI scoring was obtained using settings of “masked” and “500 nt” as the maximum distance.

### Transfection, cDNA synthesis, Sanger sequencing

The reference and variant RHCglo plasmids were transfected into the HEK293FT cells using the Lipofectamine 2000 reagent (Thermo Fisher Scientific) using a suspension transfection method as described previously^7^. Twenty-four hours after transfection, cells were lysed and total RNA extracted using an automated nucleic acid extraction instrument without DNase I treatment (Xtract16, Autogen). Fifty ng of RNA samples were used for the 1^st^ strand cDNA synthesis in 10 μL reactions using the iScript Reverse Transcription Supermix (Bio-Rad). PCR amplicons were visualized on E-Gel precast agarose gels (Thermo Fisher Scientific). PCR and Sanger sequencing were performed as previously described^7^.

### qPCR and dPCR

The qPCR reactions were performed using the Luna Universal Probe qPCR Master Mix (NEB) in duplicates using primers qMini-M and qMini-P on a QuantStudio 3 Real-Time PCR system (Thermo Fisher Scientific). The PCR program was 95°C for 20 sec, and 40 cycles of 95 °C for 15 sec and 60 °C for 40 sec. The Crossing threshold (Ct) values for qMini-M and -P were set at 0.2 ΔRn with the *RPP30* gene used for normalization in the qPCR. After qPCR, the cDNA sample was diluted to the equivalent concentration of Ct 26 for dPCR if its Ct was lower than 26 using the ΔCt method. The ddPCR supermix for Probes (Bio-Rad) was used for dPCR using the QX200 Droplet Digital PCR System (Bio-Rad). The cycling condition for dPCR was: 95°C for 10 min, and 40 cycles of 94°C for 30 sec and 60°C for 1 min, followed by 98°C for 10 min then hold at 4°C.

## Data Availability

All data produced in the present work are contained in the manuscript.

## Acknowledgements

This research was supported by the Intramural Research Program of the NIH, National Eye Institute.

## Legends

**Extended Data Fig. 1.**
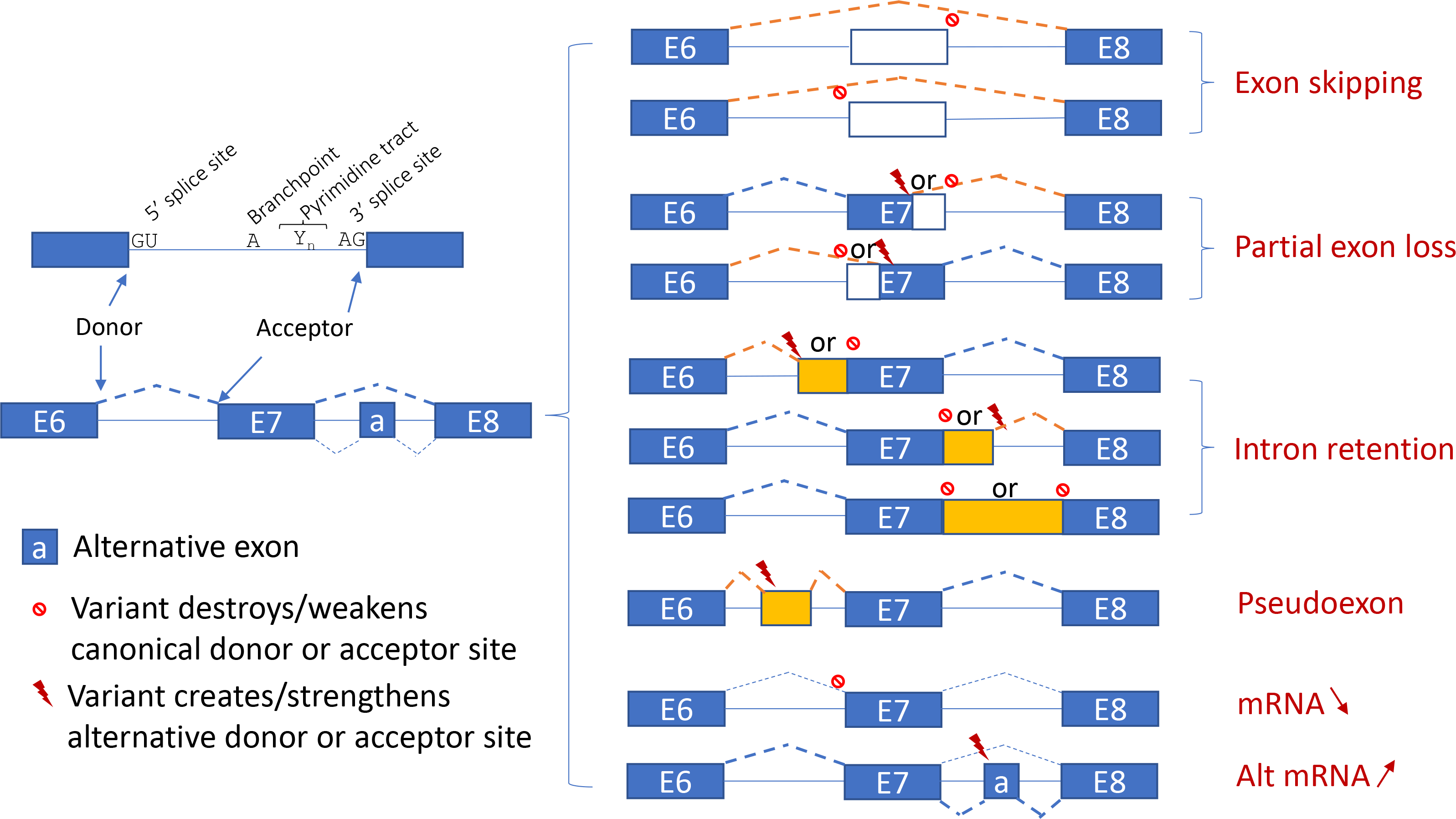
The splicing outcome diagram. The donor and acceptor dinucleotides GU-AG, branchpoints, and the pyrimidine tract found upstream of the 3’ splice site play crucial roles in the process of pre-mRNA splicing by interacting with components in the spliceosome. Splicing could occur at different efficiencies among splicing sites to produce various amounts of transcript isoforms as illustrated by dashed lines with different weights. Depending on the local sequence context, variants may change mature mRNA sequences by using alternative donor or acceptor sites, may alter mature mRNA levels by affecting splicing efficiencies, or may affect both mRNA sequences and levels.

**Extended Data Fig. 2.**
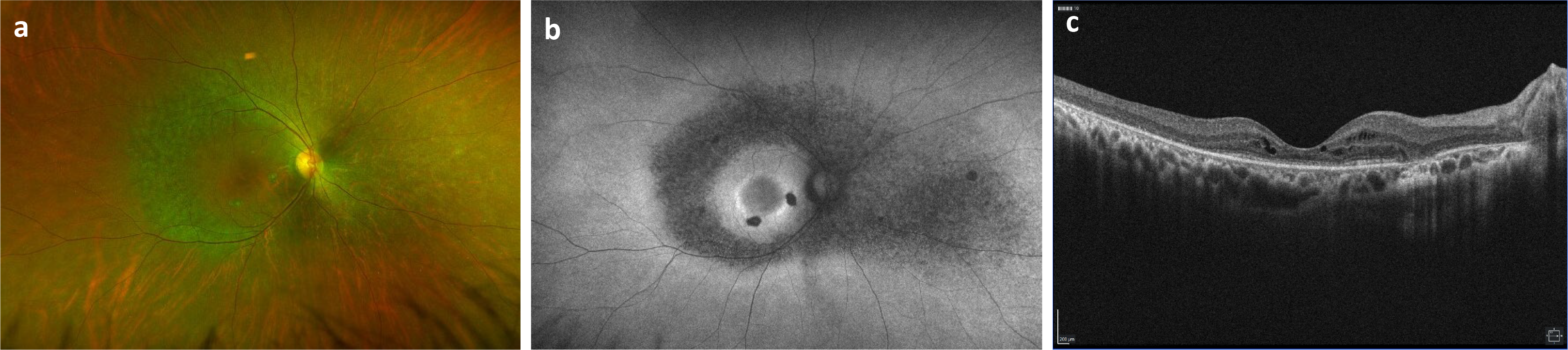
Clinical retinal imaging of the right eye of the patient with *PRPF8* c.5506-11_5506-10del variant. **a)** Wide-field color fundus image shows vascular attenuation and retinal pigment epithelial granularity along the arcades and nasally, highlighted further with fundus autofluorescence (FAF) imaging (**b**). FAF imaging also shows areas of discreet atrophy and a subtle hyper-autofluorescent around the macula. **c)** Optical coherence tomography demonstrates characteristic findings of a rod-cone dystrophy, including loss of photoreceptors and macular cystic changes.

**Extended Data Fig. 3.**
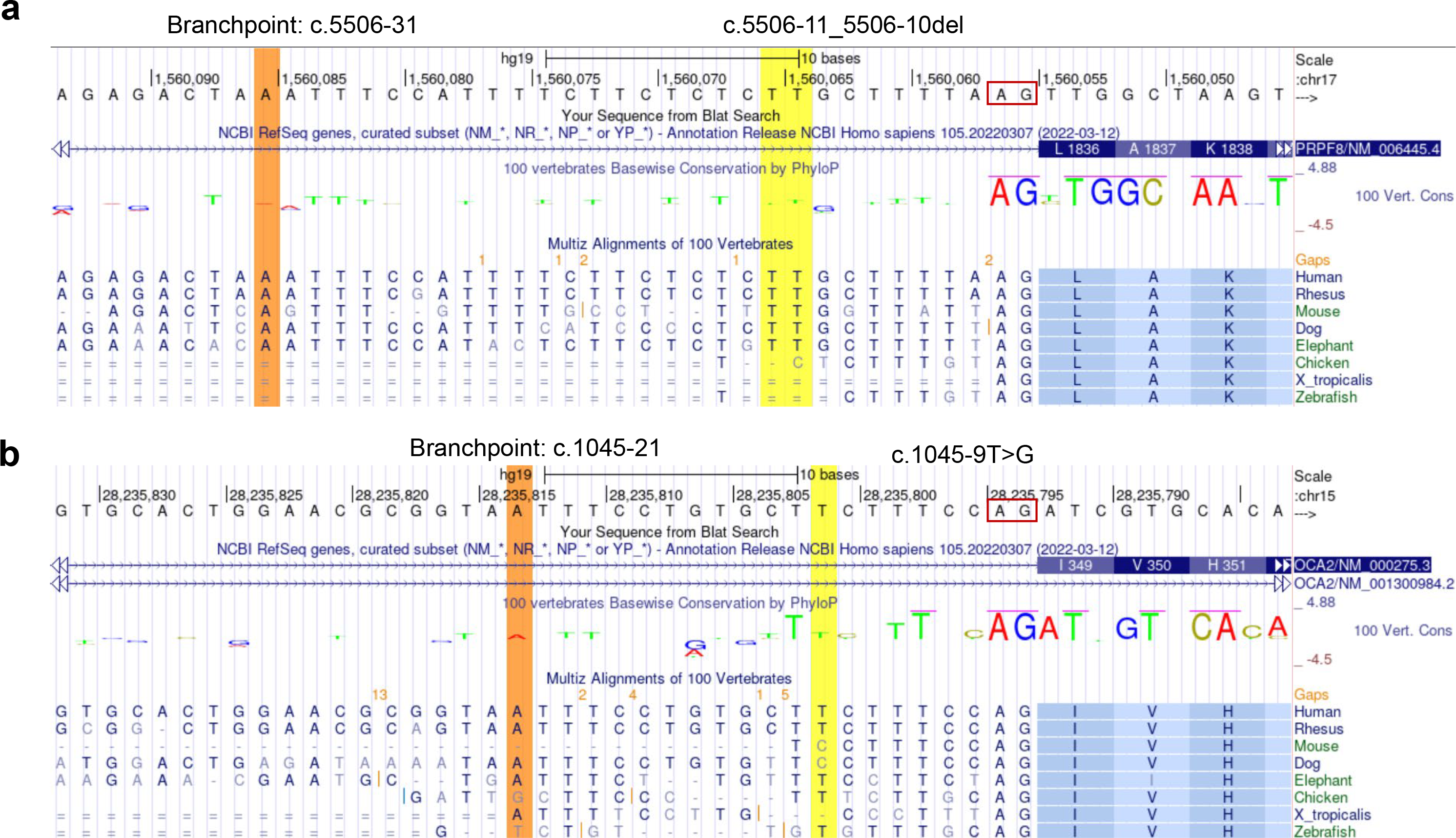
The sequence and conservation of two variants in the polypyrimidine tract. **a)** The *PRPF8* c.5506-11_5506-10del variant deletes two TT (UU) dinucleotides from the 26 nt polypyrimidine tract. **b)** The *OCA2* c.1045-9T>G mutates one T from the 15 nt polypyrimidine tract. The predicted branchpoint point (orange) was obtained using the LaBranchoR tool^14^. Red box, acceptor site. The UCSC genome browser of the hg19 version accessed on 10/23/2023. Note that both genes are on the minus strand and the genome browser view was shown in the reverse mode.

**Supplementary Table 1. Summary of participants and variant minigene assay results**

**Supplementary Table 2. Primers and probes**

**Supplementary Table 3. Summary of literature review for subjects with the c.11+5G>A variant in *RPE65***

